# Estimated population-level impact of pneumococcal conjugate vaccines against all-cause pneumonia mortality among unvaccinated age groups in five Latin American countries

**DOI:** 10.1101/2023.08.08.23293814

**Authors:** Ottavia Prunas, Kayoko Shioda, Cristiana M. Toscano, Magdalena Bastias, Maria Teresa Valenzuela-Bravo, Janepsy Diaz Tito, Joshua L. Warren, Daniel M. Weinberger, Lucia H. de Oliveira

## Abstract

**Background:** Pneumococcal conjugate vaccines (PCVs) provide strong direct protection in children, while limited data are available on their indirect effect on mortality among older age groups. This multi-country study aimed to assess the population-level impact of pediatric PCVs on all-cause pneumonia mortality among ≥5 years of age, and invasive pneumococcal disease (IPD) cases in Chile.

**Methods:** Demographic and mortality data from Argentina, Brazil, Chile, Colombia, and Mexico were collected for various age strata considering the ≥ 5-year-old population, from 2000-2020. IPD cases in Chile were also evaluated. Time series models were employed to evaluate changes in all-cause pneumonia deaths during the post-vaccination period, with other causes of death used as synthetic controls for unrelated temporal trends.

**Results:** No significant change in death rates due to all-cause pneumonia was detected following PCV introduction among most age groups and countries. The proportion of IPD cases caused by vaccine serotypes decreased from 29% (2012) to 6% (2022) among ≥65 years in Chile.

**Discussion:** While an effect of PCV against pneumonia deaths (a broad clinical definition) was not detected, evidence of indirect PCV impact was observed among vaccine-type-specific IPD cases.

## Introduction

Pneumococcus is among the leading causes of severe bacterial disease and death, with children <5 years of age and older adults carrying the largest burden worldwide [1, 2]. The most severe form of disease, known as invasive pneumococcal disease (IPD), can lead to pneumonia, meningitis, and septicemia [3]. Pneumococcal infections cause about 300,000 deaths in children <5 years of age globally each year, and a large, but poorly characterized, burden in adults [4].

Pneumococcal conjugate vaccines (PCVs) have now been introduced in >140 countries, including in many low- and middle-income settings, with countries in the Americas being among the first lower and middle-income countries (LMICs) to introduce pneumococcal conjugate vaccines into their national immunization programs for infants [1, 5, 6]. This widespread introduction of PCVs for use in infants resulted in a substantial decrease in the incidence of IPD due to PCV-targeted serotypes in children [7]. These vaccines also reduced transmission of pneumococcus from the upper respiratory tract [8]. Therefore, vaccination of children with PCVs resulted in indirect benefits for unvaccinated groups, especially in older adults [9, 10]. For example, after the introduction of PCV7 in 2000 in the United States, a substantial decline was observed across all age groups for IPD as well as pneumococcal pneumonia hospitalizations and deaths [11]. In South Africa, PCV introduction was associated with reductions in deaths due to all-cause pneumonia in children and teenagers but not in adults [12]. The magnitude of the reduction in rates of IPD varied depending on population demographics, the proportion of cases caused by vaccine-targeted serotypes, the degree to which the incidence of non-vaccine serotypes increased following vaccine introduction (‘serotype replacement’), and contact patterns in the population [13, 14].

There are various practical and technical challenges when estimating the population-level impact of PCVs. In many settings, particularly in LMICs, there is a lack of systematic surveillance for pneumococcal diseases. Specific bacterial or viral causes of disease or death are often not recorded in administrative records. Evaluations of PCV impact, therefore, often focus on non-specific outcomes, such as all-cause pneumonia, instead of pneumococcal pneumonia [1]. Detecting changes in all-cause pneumonia associated with introduction of PCVs is challenging because pneumococcus is just one of many etiologies of pneumonia [1], and thus, the fraction of all-cause pneumonia mortality that could be prevented by PCVs is relatively small. Furthermore, the overall impact of PCVs might be masked due to the replacement of serotypes or counterbalancing trends in mortality. Previous studies employed various statistical methods to overcome these challenges and detect changes in all-cause pneumonia in children during the post-PCV period [15, 16]. To date, nearly all countries in the Latin America and Caribbean (LAC) Region (n=25) have introduced PCV in their routine immunization schedules [17].

In this multi-country study, we aimed to investigate the population-level impact of PCVs against all-cause pneumonia mortality among people ≥5 years of age and against invasive pneumococcal disease (IPD) cases in Chile, following our previous study evaluating the impact among children <5 years of age. We used standardized data from five middle-income countries in Latin America and employed several complementary analytic approaches to overcome data challenges and increase the robustness of our findings.

## Methods

### Mortality Data

Mortality data from Argentina, Brazil, Colombia, Chile, and Mexico were obtained from national mortality registries in each country, considering all cases of death, and the following age-strata: 5-19, 20-39, 40-64, 65-79, and 80+ years. All countries conducted standardized data cleaning and quality control [1].

We extracted the month and year of birth and death for each death record, age of the individual at death, and the primary and underlying causes of death, as coded in International Classification of Diseases, Tenth Revision (ICD-10), similarly to our previous analysis [1].

### Study periods

We considered three immunization periods in our analysis: (1) pre-vaccine period; (2) transition period, defined as the first 12 months after PCV introduction to allow vaccine uptake to stabilize; and (3) evaluation or post-vaccine period, during which the impact of PCV was evaluated. Study periods for each country were defined depending on the timing of universal introduction of PCVs in each country, respectively (Table 1).

**Table 1:** Study periods in participant countries.

| **Country** | **Pre-vaccine period** | **Date of PCV introduction** | **Transition period** | **Evaluation period** |
| --- | --- | --- | --- | --- |
| Argentina | Jan 2005 - Jan 2012 | Jan 2012 | Feb 2012 - Dec 2012 | Jan 2013 - Dec 2019 |
| Brazil | Jan 2005 - Feb 2010 | Mar 2010 | Apr 2010 - Mar 2011 | Apr 2011 - Nov 2020 |
| Chile | Jan 2005 - Jan 2011 | Jan 2011 | Feb 2011 - Jan 2012 | Feb 2012 - Dec 2018 |
| Colombia | Jan 2005 - Jan 2011 | Jan 2011 | Feb 2011 - Jan 2012 | Feb 2012 - Dec 2019 |
| Mexico | Jan 1999 - Jan 2008 | Jan 2008 | Feb 2008 - Jan 2009 | Feb 2009 - Dec 2019 |

### Data on pneumococcal serotypes of IPD cases in Chile

We obtained data on pneumococcal serotypes from the national surveillance for IPD cases in Chile from 2012 (one year after the introduction of PCV) to 2022. We analyzed data for those aged ≥65 years and calculated the proportion of IPD cases caused by 1) PCV-10 serotypes, 2) serotype 3, 6A, and 19A (additional serotypes included in PCV13), and 3) non-vaccine-type serotypes for each year [18].

### Statistical approaches

The goal of our analysis was to evaluate the population-level impact of PCVs in age groups ≥5 years old following universal introduction of PCV in infants. The outcome of interest was death due to all-cause pneumonia, defined as having an ICD-10 code for either the primary or underlying cause of death in the range of J12-J18. The age group of 5-19 years will capture both direct and indirect effects in many of these countries due to the long follow-up period, while for the remaining age groups (20+ years), our analysis would capture indirect effects only.

We used synthetic control (SC) models as our primary analysis for all age groups and countries to reduce biases and to isolate changes in all-cause pneumonia mortality caused by PCV as opposed to those caused by other factors, such as changes in underlying health of the population and reporting of mortality records. This approach uses other causes of death not affected by the vaccine, to adjust for shared temporal trends. Advantages of using the SC approach have been shown previously [7]. Among the most important, the SC approach is more robust to unmeasured confounding and biases than other methods (e.g., interrupted time series, ITS) because it uses information on control conditions that were affected by common unrelated factors but not by the intervention of interest (i.e., PCV in this study). Additionally, SC models select an appropriate set of controls based on the pre-vaccine data; this step is performed in a data-driven way, without requiring users to select controls *a priori*. Finally, unlike ITS models, the SC method does not need to assume that trends in the pre-vaccine period continue linearly in the post-vaccine period or assume that changes happen at specified time points. All analyses were performed in R (Vienna, Austria).

### Synthetic control (SC) model

The version of the SC model used in this study is a negative binomial regression model where the outcome was the observed number of all-cause pneumonia deaths (ICD-10 code: J12-J18) per month. The covariates or control diseases were monthly time series for other ICD-10 chapters or subchapters of death per month that are not likely to be influenced by PCVs, including disease of the circulatory system (I00–I99), skin (L00–L99), musculoskeletal system (M00–M99), and genitourinary system (N00–N99) (see a supplementary file for the full list of control diseases). We fitted the SC model to the pre-PCV vaccine data from each age group in each country separately, using the rjags package [19] regularization prior distribution similar to those used in ridge regression was assigned for the control variable regression parameters to deal with the potentially large number of control variables under consideration and correlation between them. These selected controls were combined into a composite used to generate a counterfactual of the number of all-cause pneumonia deaths that would have occurred without the intervention. This prediction was then compared with observed all-cause pneumonia deaths during the evaluation period for each age group in each country. We quantified the impact of PCV by computing a rate ratio (RR) for each age group in each country, which is the total cumulative number of observed all-cause pneumonia deaths divided by the counterfactual cumulative number of predicted all-cause pneumonia deaths during the evaluation period. Posterior medians and 2.5th and 97.5th posterior percentiles were reported as point estimates and 95% CrIs of RR, respectively. The model used here differs from SC models used in previous studies, which used a quasi-Poisson model together with a spike-and-slab prior as the regularization prior distribution [1]. This version of the model did not converge well, leading us to explore alternative structures.

### Sensitivity analysis

In addition to the SC models, we applied a more commonly used approach, interrupted time series (ITS) models [20]. We fit a negative binomial regression model to the monthly time series data on all-cause pneumonia deaths over the entire study period, with an offset being all-cause mortality other than those caused by respiratory illness (ICD-10 J chapter). Separate models were fit for each age group in each country separately. A linear trend is included throughout the entire study period; also, we included two changes in intercept and slope to model the impact of the intervention. The first change in intercept and slope represented the *transition* period; while the second change in intercept and slope represented the *evaluation* period. The counterfactual number of all-cause pneumonia deaths was then generated by removing those slopes and intercepts. The impact of PCV was quantified based on a RR, this time using the model fitted to the observed time series of pneumonia deaths divided by the expected number of pneumonia deaths if the intervention had not been introduced for the evaluation period. As a second sensitivity model, we combined the synthetic control and interrupted time series approach together into the SC+ITS model. We included all the control variables as in the SC model and we used the ridge regression style prior distributions to provide robust estimation results. Additionally, we included two slopes and intercepts to quantify the impact of the intervention, and we fitted the model to the entire time series of data. The impact of PCV was quantified following the approach described for the ITS model.

### Ethical considerations

The study protocol was reviewed and approved by institutional review boards in each country and also by the Pan American Health Organization Ethical Research Committee (PAHOERC).

### Availability of code and data

The aggregated time series data and code can be found in the following github repository: https://github.com/weinbergerlab/Paho_project_PCV_adult

## Results

### Descriptive results

The proportion of all deaths caused by all-cause pneumonia (ICD-10 code J12-J18 as the primary cause of death) varied by country and age group (Figure 1). In general, the proportion was highest among the oldest age groups (65-80, 80+ years), ranging from 5% to 10%, and lowest among younger age groups, ranging from 1% to 5%. The proportion gradually increased in some age groups in some countries, especially older age groups, (e.g., Argentina and Brazil).

**Figure 1:**
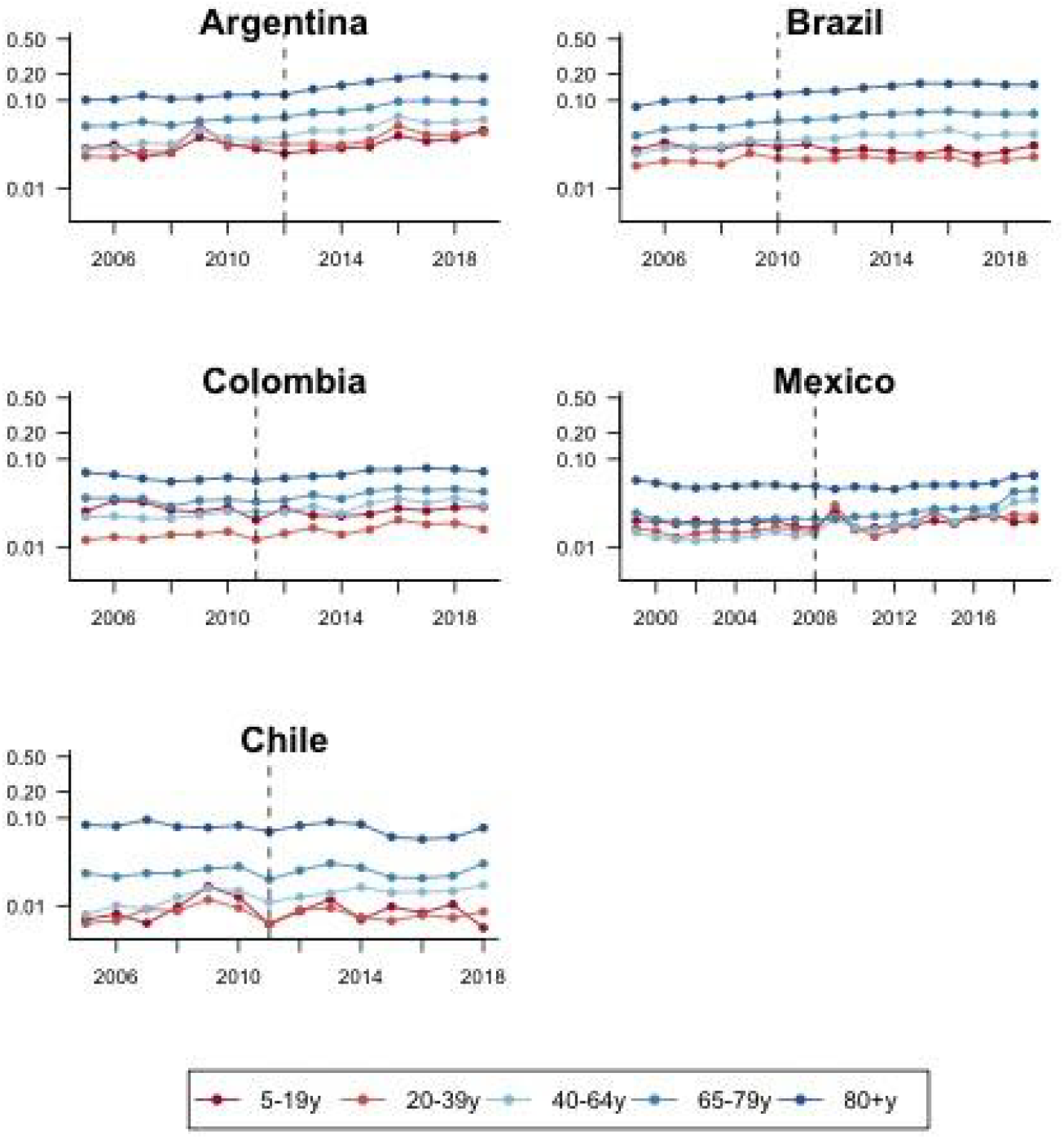
Annual time series for the proportion of all deaths recorded as pneumonia (International Classification of Diseases, Tenth Revision [ICD-10] codes J12–J18) as primary cause of death, by age group in five Latin American countries. Vertical dashed lines represent the timing of universal pneumococcal conjugate (PCV) vaccine introduction.

### Changes in all-cause pneumonia mortality across age groups and countries

In general, there was not a detectable change in rates of death due to all-cause pneumonia following the introduction of PCV among most age groups in the majority of the countries (Figure 2, left panel, and Figures S1 to S5 in the supplementary file). The estimates of the RRs were generally close to one, or with wide 95% credible intervals, suggesting that there was no detectable difference between the observed and expected number of all-cause pneumonia deaths during the evaluation period, and/or that the uncertainty in the estimates was too large to draw firm conclusions.

**Figure 2:**
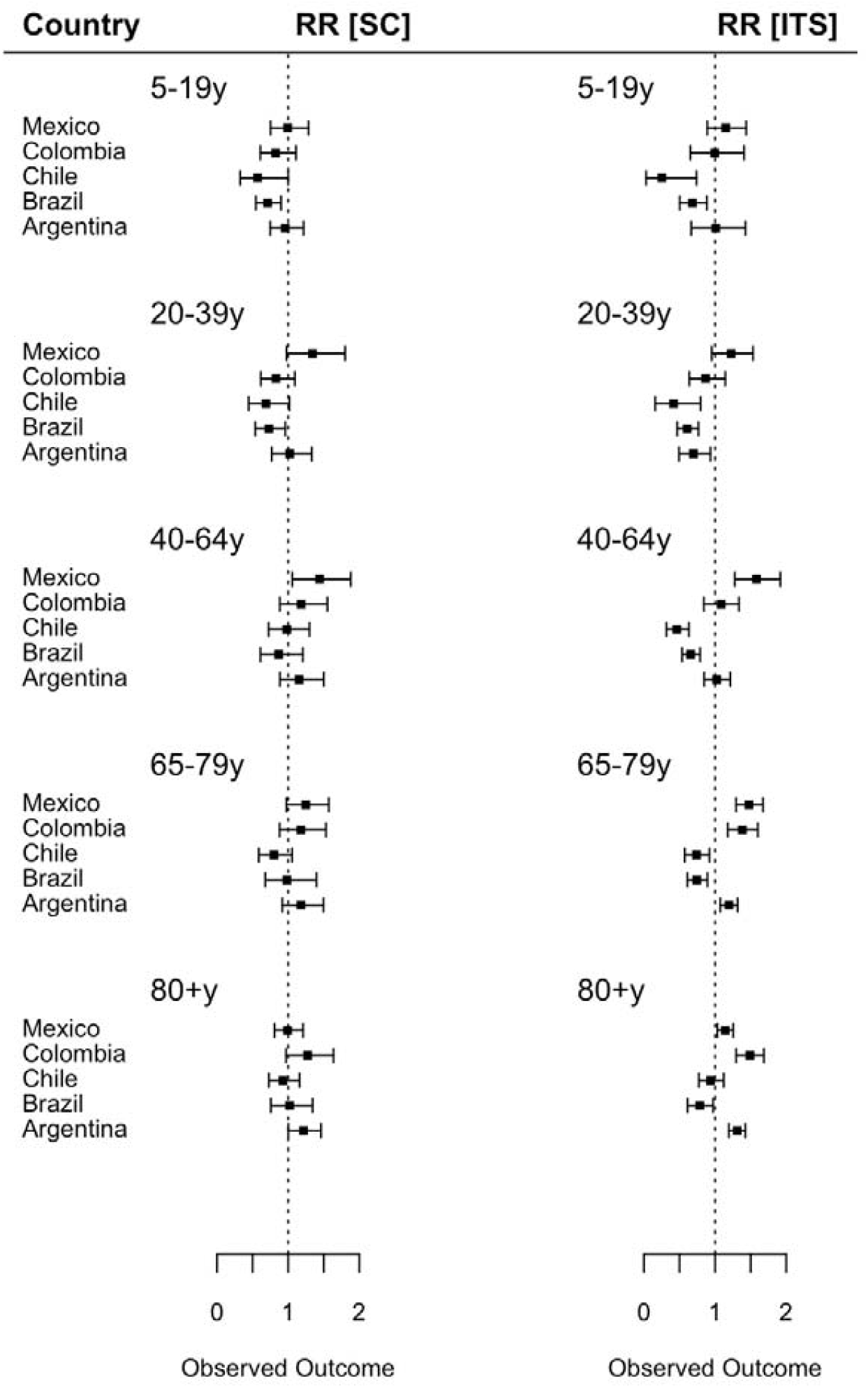
Estimated impact of pneumococcal conjugate vaccine among age groups 5-19y, 20-39y, 40-64y, 65-79y, and 80+y using the SC and ITS modeling approaches, by country. Rate ratios were calculated by dividing the cumulative number of observed pneumonia deaths by the cumulative number of predicted pneumonia deaths during the evaluation period. Black squares represent the point estimates of rate ratio and bars represent their 95% credible intervals. Abbreviation: CrI, credible interval.

Changes in all-cause pneumonia deaths in the evaluation period were observed in a few age groups in certain countries. Adults aged 40-64 years in Mexico observed an estimated increase in all-cause pneumonia mortality of 44% (RR: 1.44; 95% CrI: 1.08-1.90), with increases among those aged ≥80 years (RR: 1.21; 95% CrI: 1.00-1.46) in Argentina (Figure 2 left panel, and Figures S1 to S5 in the supplementary file). A predicted decline in all-cause pneumonia mortality was observed in Brazil in age groups 5-19 and 20-39 years old (RR: 0.71; 95% CrI: 0.55-0.90 and RR: 0.73; 95% CrI: 0.54-0.96).

### Distribution of pneumococcus serotype in IPD cases in Chile

Among IPD cases aged ≥65 years in Chile, we found that vaccine-type pneumococcus, especially those included in PCV10, decreased every year after PCV introduction ranging from 29% in 2012 to 6% in 2022 (Figure 3).

**Figure 3:**
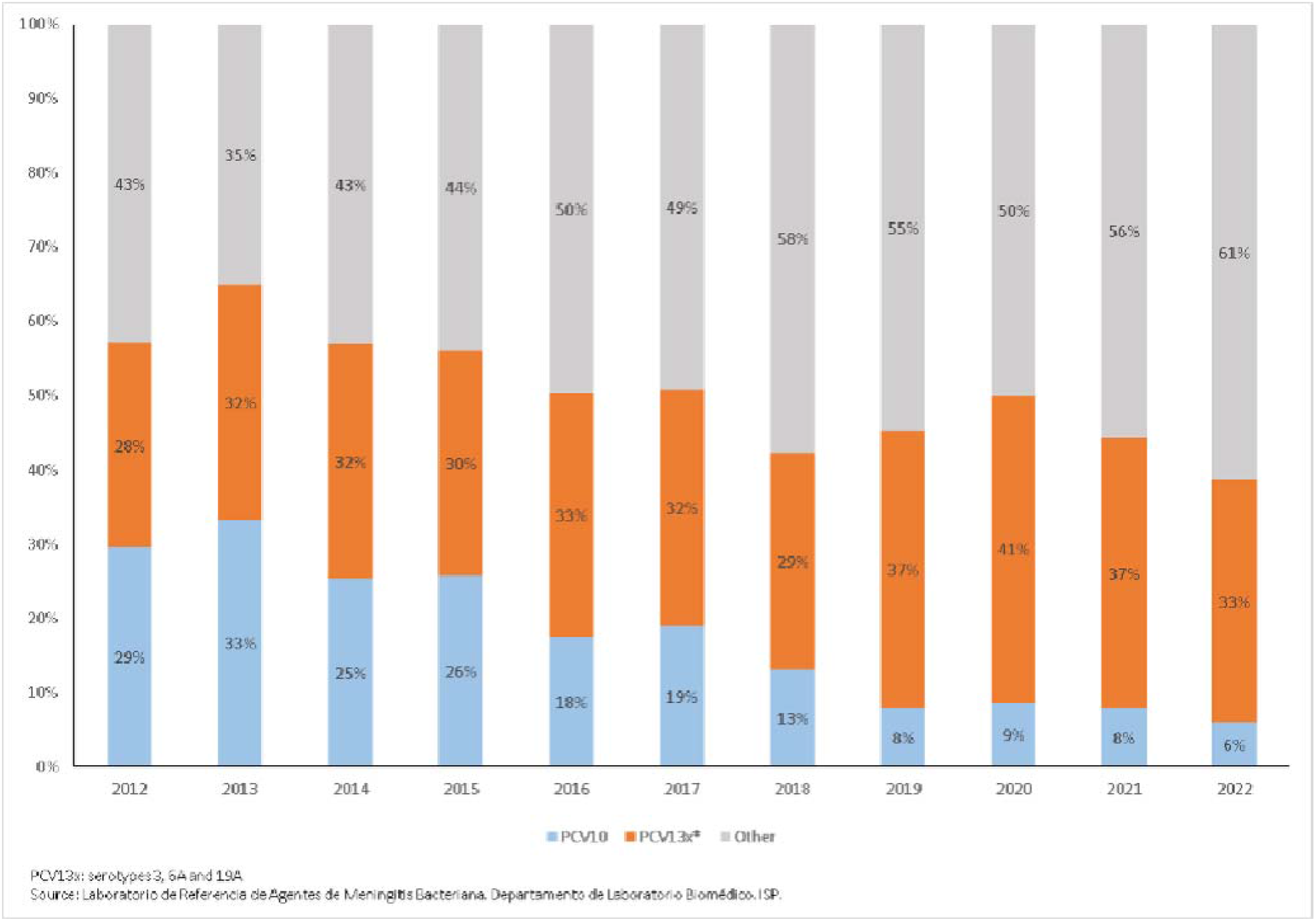
Serotype distribution of *Streptococcus pneumoniae* strains isolated from Invasive Pneumococcal Disease (IPD) among older adults ≥65 years of age. Chile, 2012-2022.

### Sensitivity analyses

Changes in all-cause pneumonia mortality following the introduction of PCV estimated by the ITS approach varied across age groups and countries (Figure 2 and Figures S6 to S10). RRs estimated by the ITS approach were qualitatively different from those estimated by the SC approach, with estimated declines or increases in some age groups and countries, where a change was not estimated with the SC method. For example, the ITS approach predicted an estimated decline in deaths in all age groups ≤ 79 years in Brazil and Chile (∼25% to 80%). The SC+ITS results were in line with the ITS model in most age groups and countries with a few exceptions (Figure S11). In Argentina, the SC+ITS seemed to correctly adjust for the bias among those aged 64-79 years (RR: 1.04 with 95% CrI 0.83, 1.25). Also, the RRs from the SC+ITS were close to 1 in a few age groups in Brazil and Chile, confirming the SC model’s results (Figure S11).

## Discussion

This multi-country study represents the first comprehensive, multinational effort to measure the indirect effects of PCV on all-cause pneumonia mortality across wide age ranges in the Latin American region. Our analyses relied on national mortality registries in each country and used different methodological approaches, finding no consistent evidence of a decline in all-cause pneumonia deaths among individuals ≥5 years of age following PCV introduction in most settings. In our previous study where we employed similar approaches to evaluate the impact of PCVs among children ≤5 years of age in 10 Latin American and Caribbean countries, we found detectable declines in all-cause pneumonia deaths in Colombia (RR: 0.76; 95% CrI: 0.65-0.97) and Mexico (RR: 0.89; 95% CrI: 0.82-0.97) but not in Argentina (RR: 0.92; 95% CrI: 0.74-1.11) nor in Brazil (RR: 0.98; 95% CrI: 0.92-1.04) (Chile was not included) [1]. Given that the impact of PCV against all-cause pneumonia mortality was modest or null among directly vaccinated age groups, it was not surprising to see no impact among individuals ≥5 years of age.

Several studies have shown evidence of indirect effects of pediatric vaccination against invasive pneumococcal disease and morbidity due to pneumonia in older age groups [11, 21]. Furthermore, real-world evidence of the direct impact of PCVs on children’s mortality are largely from countries with low infant and child mortality, with a few exceptions [1, 12, 22]. In contrast, investigations into the indirect effects of PCV on pneumonia mortality in older age groups in middle-income countries remain limited [12, 22]. Our study investigates the indirect effects of PCV on pneumonia mortality in unvaccinated age groups across five middle-income countries. We believe that one of the strengths of our analyses lies in testing several methods with different assumptions about the underlying trends in the data.

The SC models estimated that all-cause pneumonia mortality increased compared to the counterfactual scenario among older age groups in Argentina and Mexico, suggesting that the PCVs had negative impact, an implausible result. There are potential factors contributing to this result. We observed an abrupt increase in all-cause pneumonia deaths during the post-PCV period among older adults in Argentina and Mexico. The SC models were unable to fully account for such increasing trends in all-cause pneumonia deaths unrelated to vaccination, likely because the changes in data trends affected respiratory diseases but did not impact control diseases. These trends were likely artifactual and could have various explanations. For example, causes of death in many settings were inconsistently reported, and there was an overuse of the codes for “unspecified” cause of death [1]. Accuracy of reporting likely improved after the H1N1 pandemic, for example in Colombia, with an improvement in the diagnosis of respiratory diseases precision thus leading to an apparent increase in trends. Conversely, a further result of the H1N1 pandemic was the overuse of classifying symptoms compatible with pneumonia immediately as such. These explanations could likely have impacted Argentina and Mexico as well. The data quality influences the robustness of the results, therefore our results should be interpreted with caution. It is generally challenging to estimate the impact of PCV against all-cause pneumonia because vaccine-type pneumococcus is only a part of the various pathogens causing pneumonia and non-vaccine-type pneumococcus may increase after the reduction of vaccine-type post PCV introduction. We found that, among IPD cases in Chile, vaccine-type pneumococcus decreased every year after PCV introduction suggesting an indirect effect of the vaccine, while non-vaccine-type pneumococcus increased among older age groups (Figure 3). Serotype replacement could have occurred in Argentina as well, which could explain why the estimated RRs were greater than 1 in older age groups.

The SC model used in this study faced challenges in accurately accounting for non-linear underlying trends in mortality data over time, particularly in certain age groups and countries. In Argentina and Mexico, for instance, the observed increase in specific age groups can be attributed to post-vaccine introduction changes that were not adequately captured by the control diseases, highlighting the limitations of the SC model in those settings. Furthermore, traditional approaches such as ITS also exhibited limitations in addressing non-linear underlying trends in various instances, resulting in counterintuitive findings among older adults in Argentina, Colombia, and Mexico (Figures S6 to S10 in the supplementary file). The ITS predicted a positive impact in Chile and Brazil across the majority of age groups. However, these results could be influenced by the linear trend assumption, potentially introducing biases (Figures S6 to S10 in the supplementary file). To mitigate these biases, we explored the performance of SC+ITS models, which demonstrated the ability to adjust for these limitations in certain settings. This suggests that it might be possible to address these biases through modifications to the existing methods. However, further investigation is required.

In conclusion, we evaluated the indirect effects of PCV on pneumonia mortality among older age groups using routinely collected data from national mortality registries in five Latin American countries. The overall evidence suggested no detectable effect of pediatric PCV vaccination on all-cause pneumonia mortality in the unvaccinated population with consistent results across most age groups and countries, although the data quality influenced the robustness of our results in a few settings. On the contrary, our analysis of IPD suggests that PCVs are indeed reducing exposure to vaccine-type pneumococci in older adults. Our findings highlight the importance of using different analytical methods to increase the robustness of the results and to detect biases in the data. Further investigation is needed to understand more robustly the indirect impact of the PCV vaccine on mortality in older adults.

## Data Availability

https://github.com/weinbergerlab/Paho_project_PCV_adult

## Acknowledgment

The authors thank the country teams and EPI programs for data collection and cleaning. The authors also thank Keith Klugman and Gail Rodgers for their critical feedback. Finally, the authors thank Rodrigo Puentes for their support with the invasive pneumococcal disease (IPD) data from Chile.

