## Supplementary file for "Estimated population-level impact of pneumococcal conjugate vaccines against all-cause pneumonia mortality among unvaccinated age groups in five Latin American countries"

Figure S1: Annual time series for the observed and counterfactual number of all-cause pneumonia deaths among individuals aged 5-19 years old in 5 Latin American countries. Dots represent the observed number of all-cause pneumonia deaths (*International Classification of Diseases, Tenth Revision* codes J12–J18). Lines and gray-shaded areas represent point estimates and 95% credible intervals of the counterfactual all-cause pneumonia deaths from the synthetic control (SC) model in the absence of pneumococcal conjugave vaccines, respectively. Vertical dashed lines show the timing of pneumococcal conjugate vaccine introduction in each country.

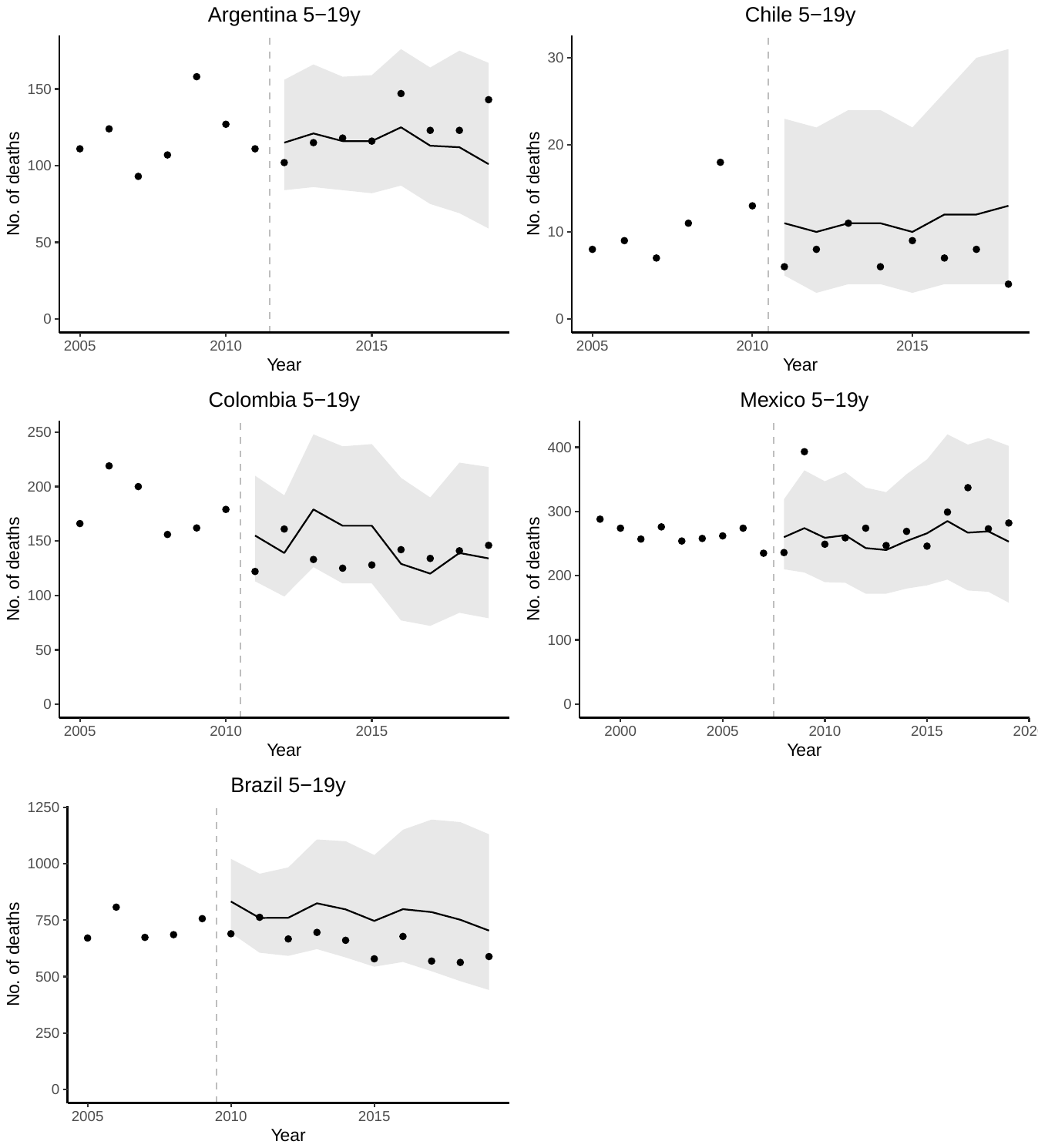

Figure S2: Annual time series for the observed and counterfactual number of pneumonia deaths among children and young adults aged 20-39 years old in 5 Latin American countries. Dots represent the observed number of pneumonia deaths (*International Classification of Diseases, Tenth Revision* codes J12–J18). Lines and gray-shaded areas represent point estimates and 95% credible intervals of the predicted pneumonia deaths from the SC model, respectively. Vertical dashed lines show the timing of pneumococcal conjugate vaccine introduction in each country.

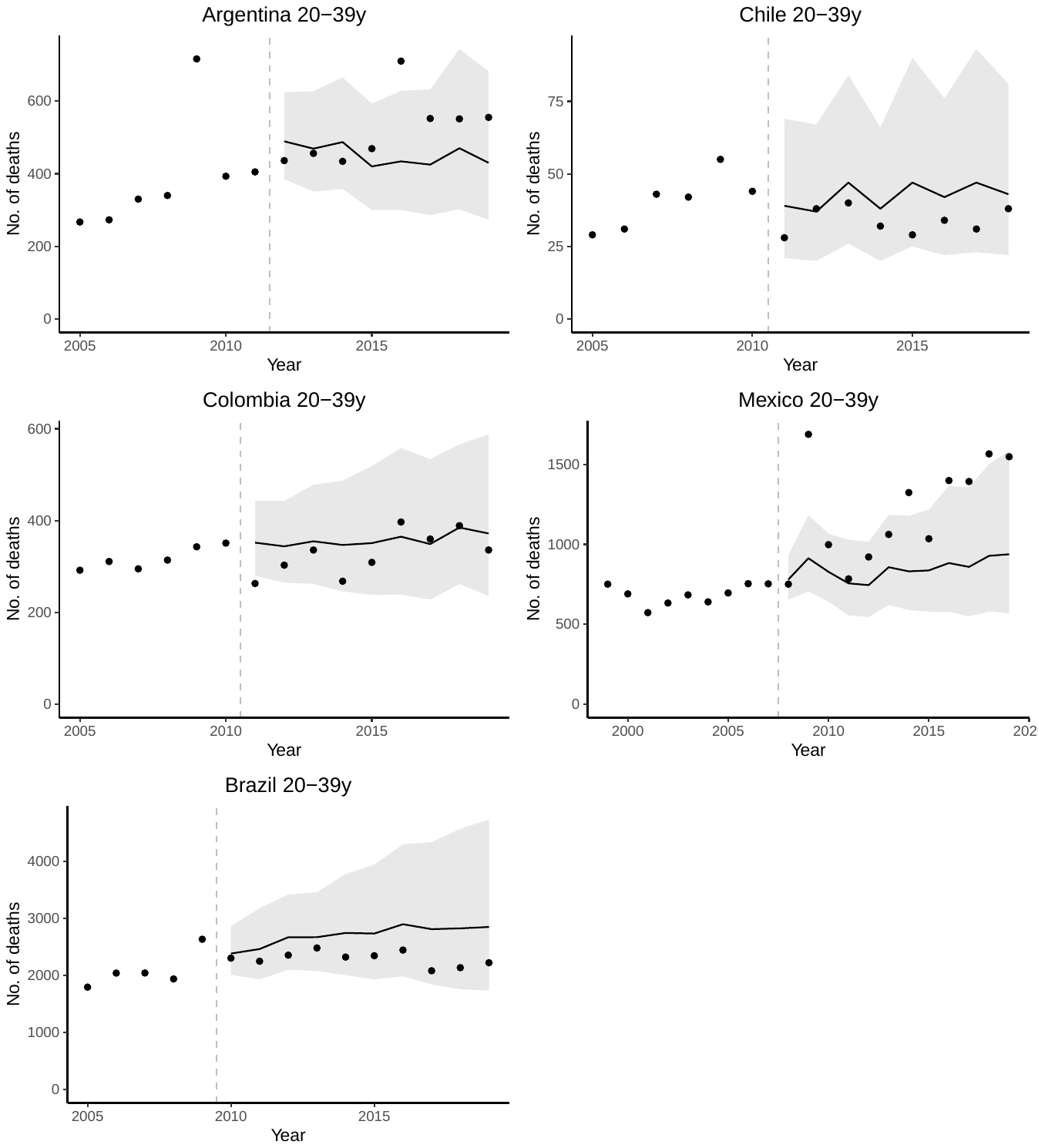

Figure S3: Annual time series for the observed and counterfactual number of pneumonia deaths among children and young adults aged 40-64 years old in 5 Latin American countries. Dots represent the observed number of pneumonia deaths (*International Classification of Diseases, Tenth Revision* codes J12–J18). Lines and gray-shaded areas represent point estimates and 95% credible intervals of the predicted pneumonia deaths from the SC model, respectively. Vertical dashed lines show the timing of pneumococcal conjugate vaccine introduction in each country.

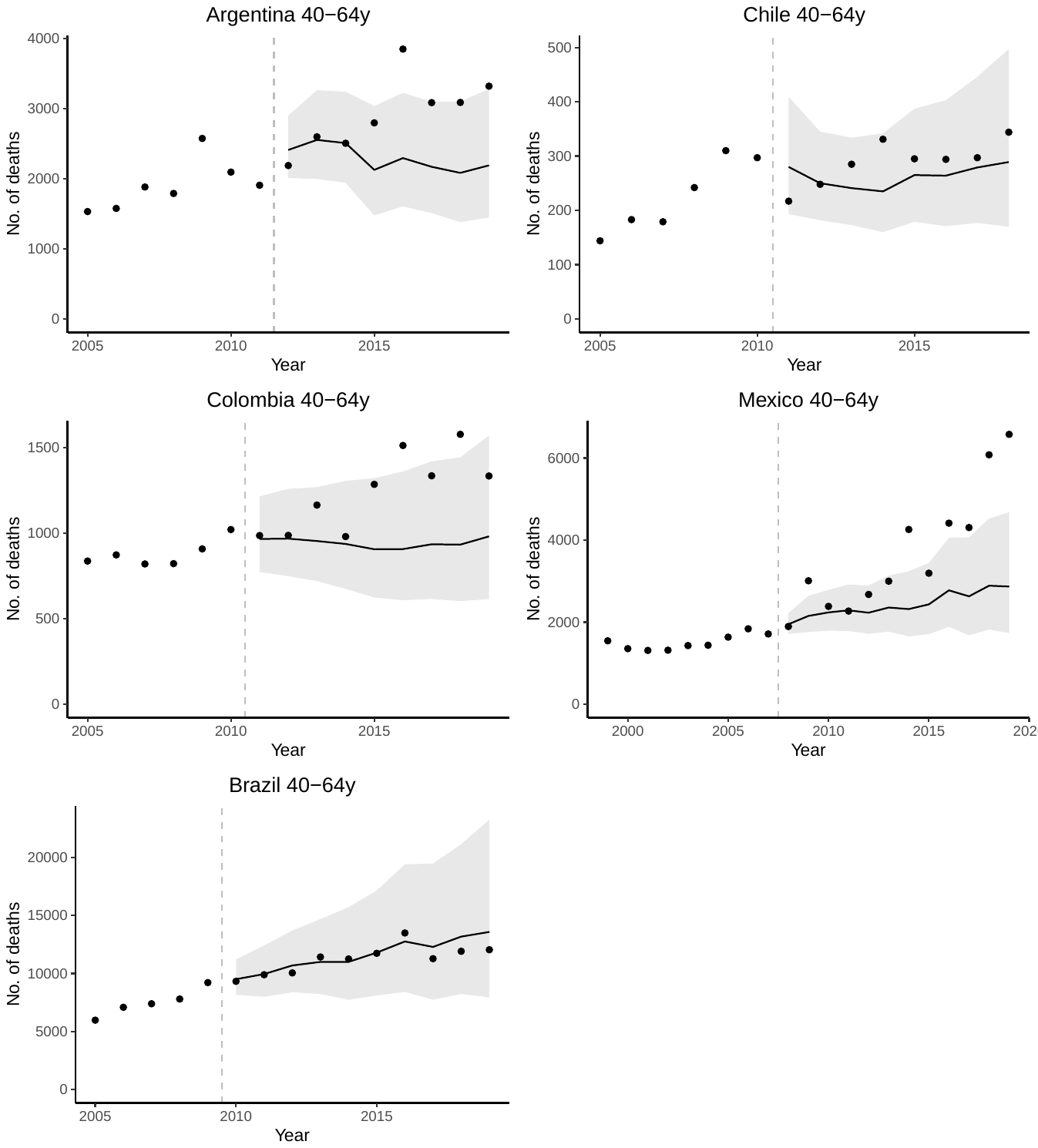

Figure S4: Annual time series for the observed and counterfactual number of pneumonia deaths among children and young adults aged 65-79 years old in 5 Latin American countries. Dots represent the observed number of pneumonia deaths (*International Classification of Diseases, Tenth Revision* codes J12–J18). Lines and gray-shaded areas represent point estimates and 95% credible intervals of the predicted pneumonia deaths from the SC model, respectively. Vertical dashed lines show the timing of pneumococcal conjugate vaccine introduction in each country.

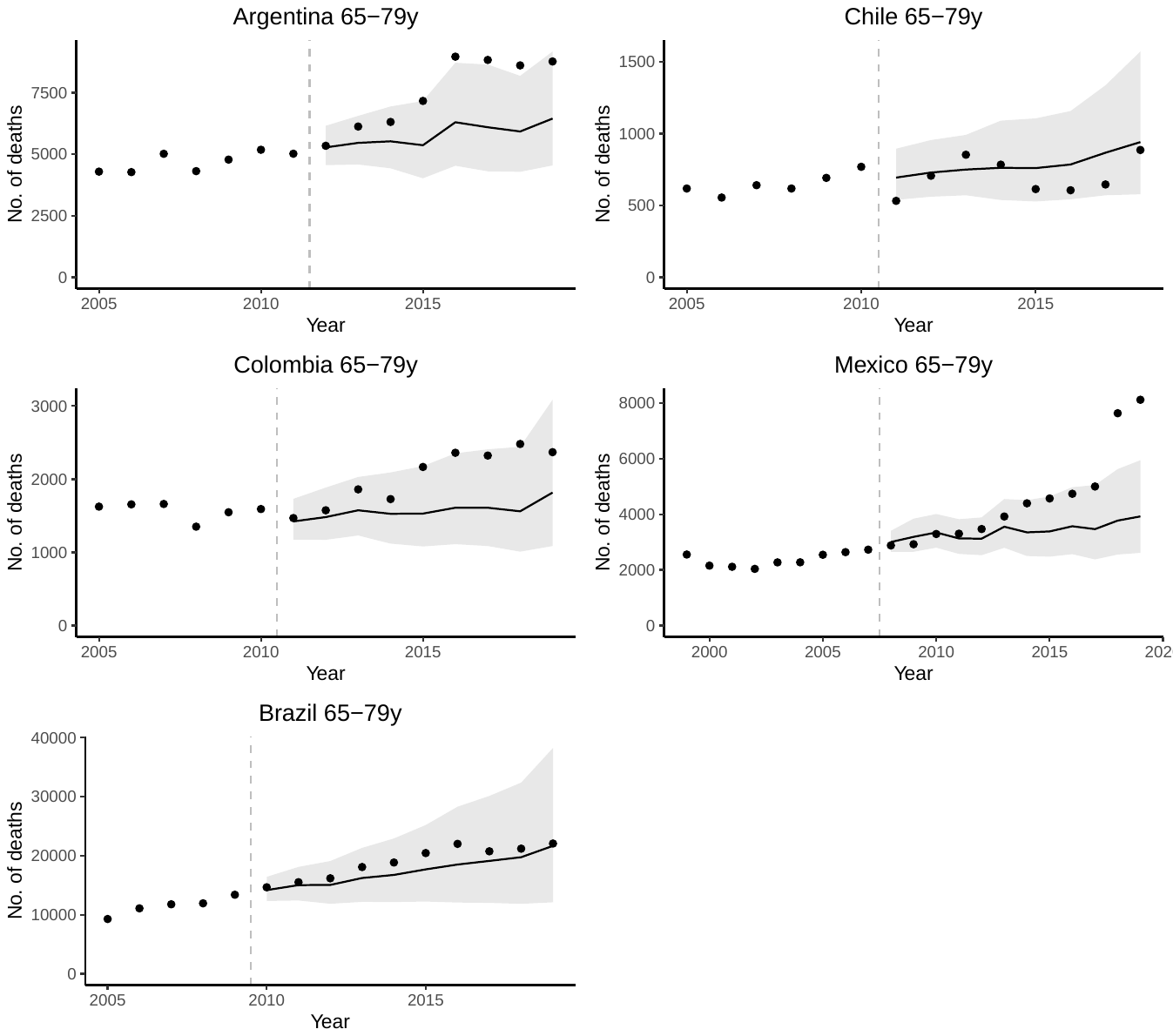

Figure S5: Annual time series for the observed and counterfactual number of pneumonia deaths among children and young adults aged 80+ years old in 5 Latin American countries. Dots represent the observed number of pneumonia deaths (*International Classification of Diseases, Tenth Revision* codes J12–J18). Lines and gray-shaded areas represent point estimates and 95% credible intervals of the predicted pneumonia deaths from the SC model, respectively. Vertical dashed lines show the timing of pneumococcal conjugate vaccine introduction in each country.

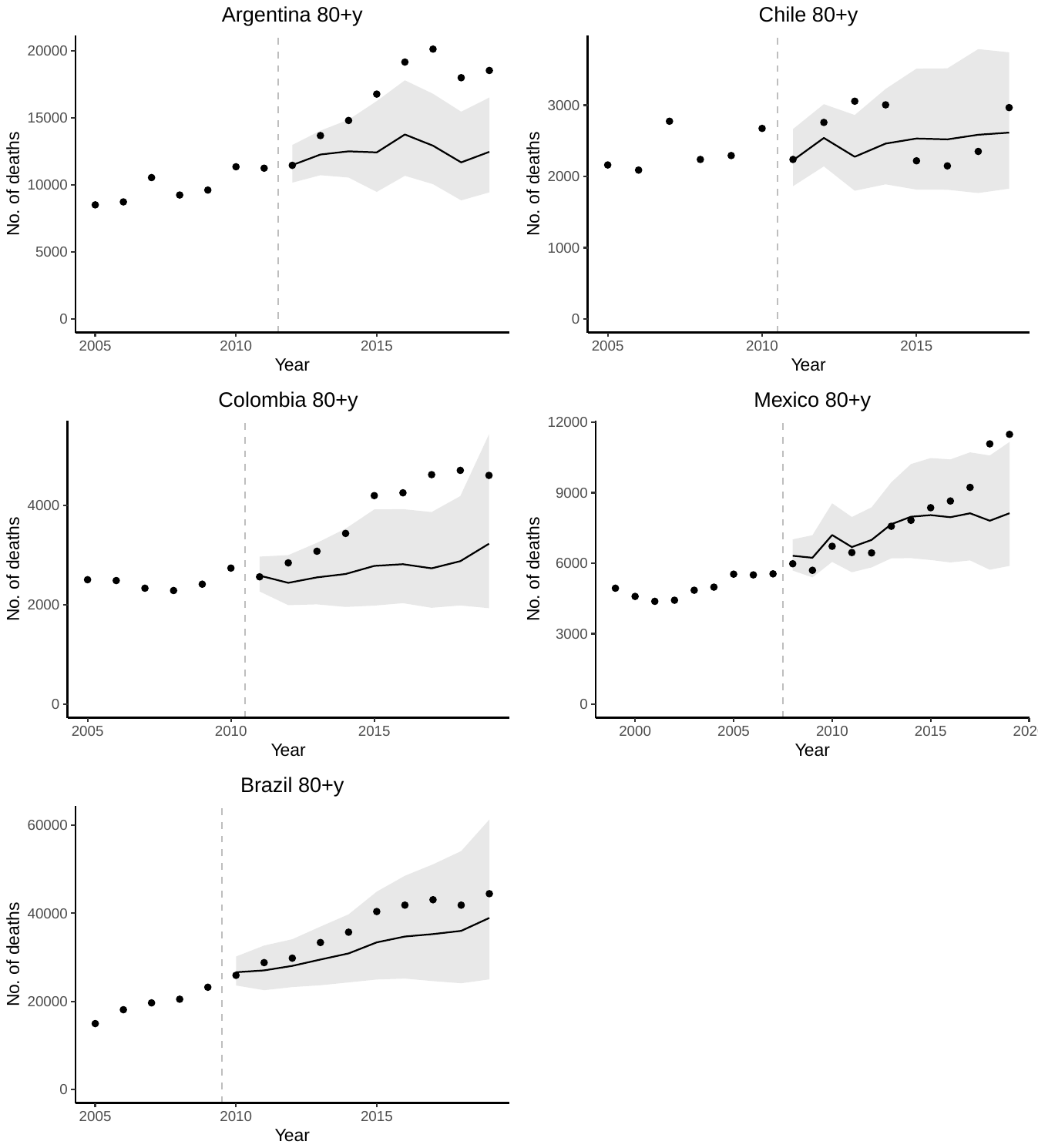

Figure S6: Annual time series for the observed and counterfactual number of all-cause pneumonia deaths among individuals aged 5-19 years old in 5 Latin American countries. Dots represent the observed number of all-cause pneumonia deaths (*International Classification of Diseases, Tenth Revision* codes J12–J18). Lines and gray-shaded areas represent point estimates and 95% credible intervals of the fitted all-cause pneumonia deaths from the interrupted time series (ITS) model, respectively. Red lines and shaded areas represent point estimates and 95% credible intervals of the counterfactual all-cause pneumonia deaths in the absence of pneumococcal conjugave vaccines, respectively. Vertical dashed lines show the timing of pneumococcal conjugate vaccine introduction in each country.

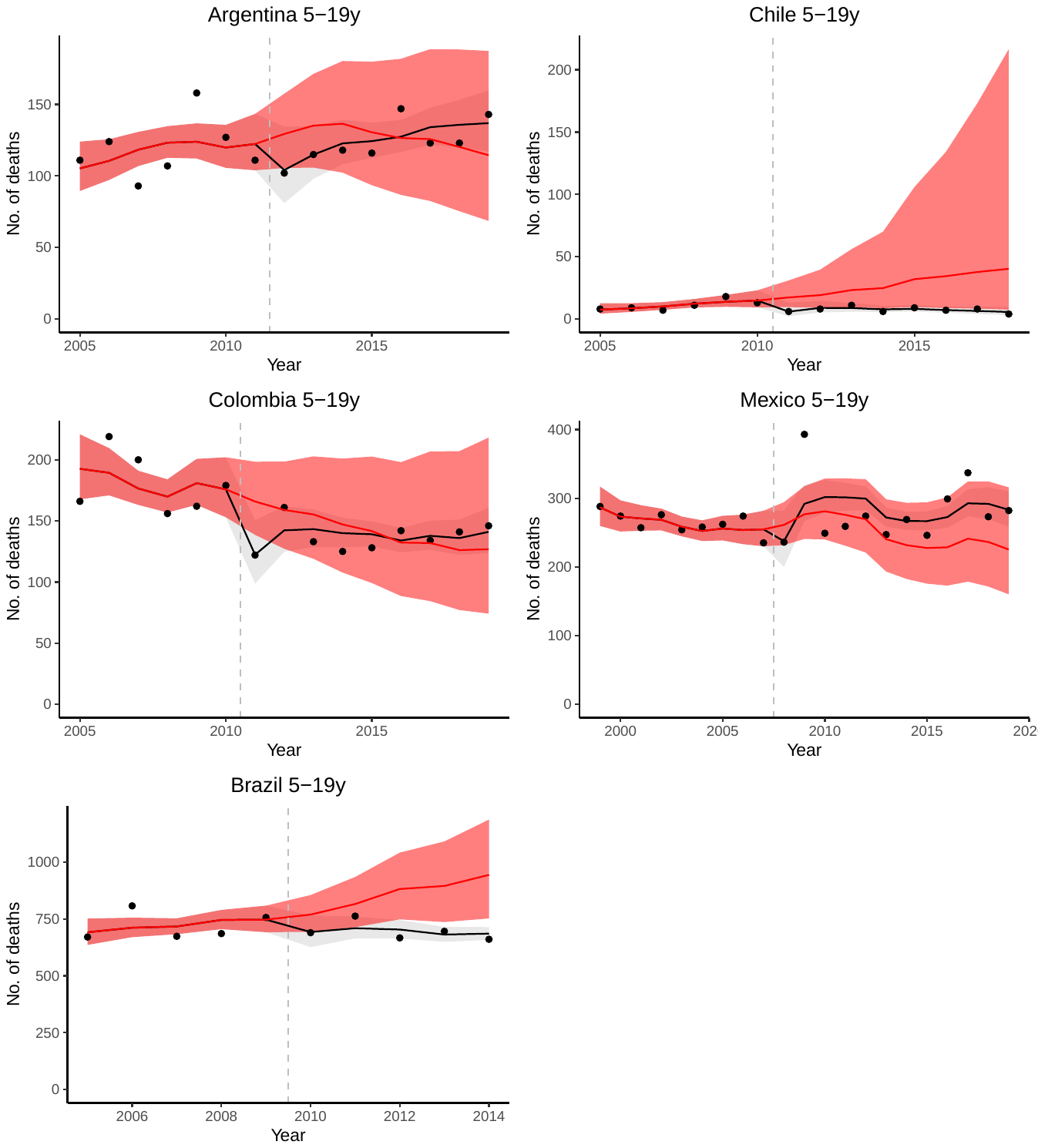

Figure S7: Annual time series for the observed and counterfactual number of pneumonia deaths among children and young adults aged 20-39 years old in 5 Latin American countries. Dots represent the observed number of pneumonia deaths (*International Classification of Diseases, Tenth Revision* codes J12–J18). Lines and gray-shaded areas represent point estimates and 95% credible intervals of the fitted pneumonia deaths from the ITS model, respectively. Red lines and shaded areas represent point estimates and 95% credible intervals of the predicted pneumonia deaths, respectively. Vertical dashed lines show the timing of pneumococcal conjugate vaccine introduction in each country.

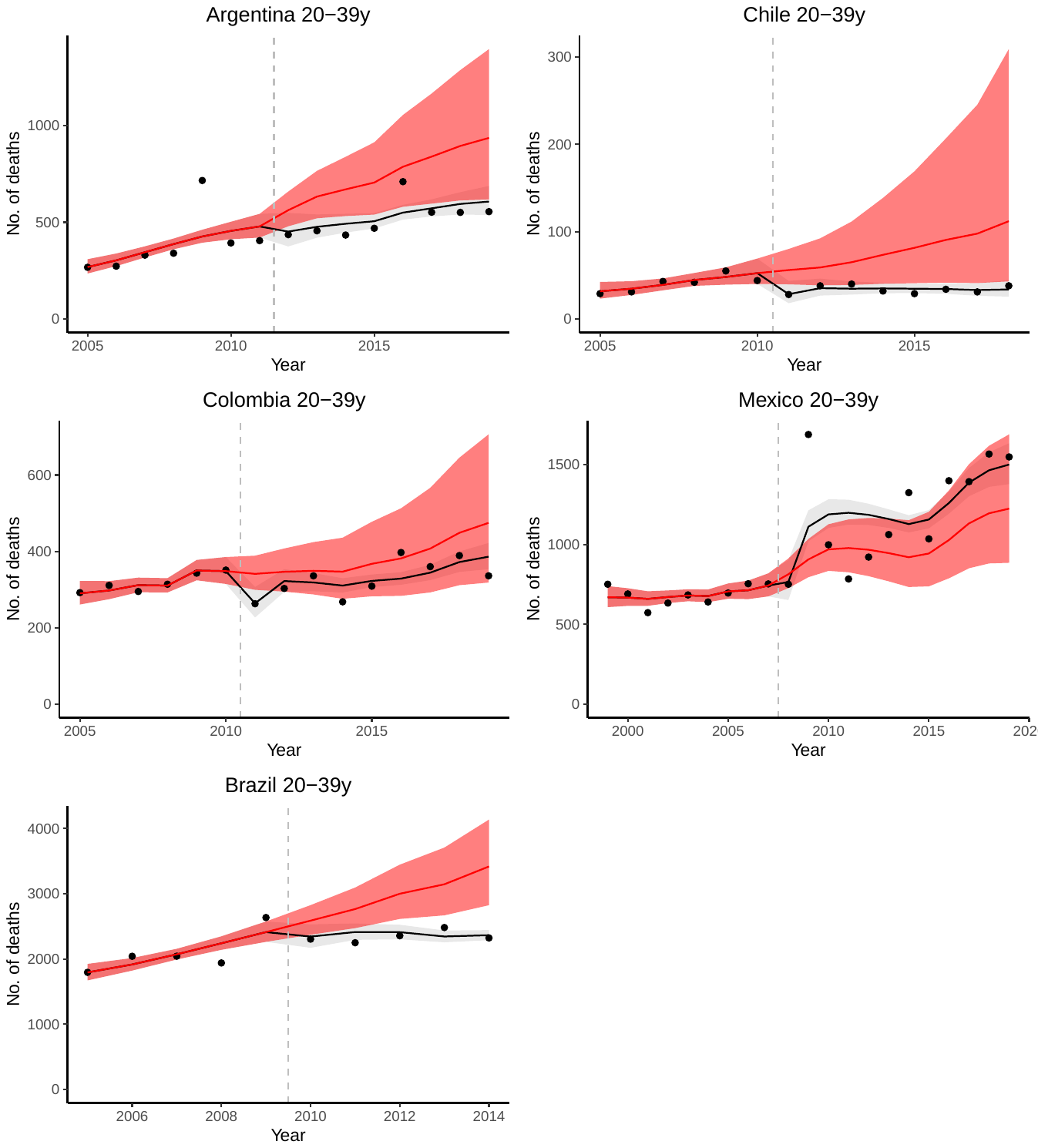

Figure S8: Annual time series for the observed and counterfactual number of pneumonia deaths among children and young adults aged 40-64 years old in 5 Latin American countries. Dots represent the observed number of pneumonia deaths (*International Classification of Diseases, Tenth Revision* codes J12–J18). Lines and gray-shaded areas represent point estimates and 95% credible intervals of the fitted pneumonia deaths from the ITS model, respectively. Red lines and shaded areas represent point estimates and 95% credible intervals of the predicted pneumonia deaths, respectively. Vertical dashed lines show the timing of pneumococcal conjugate vaccine introduction in each country.

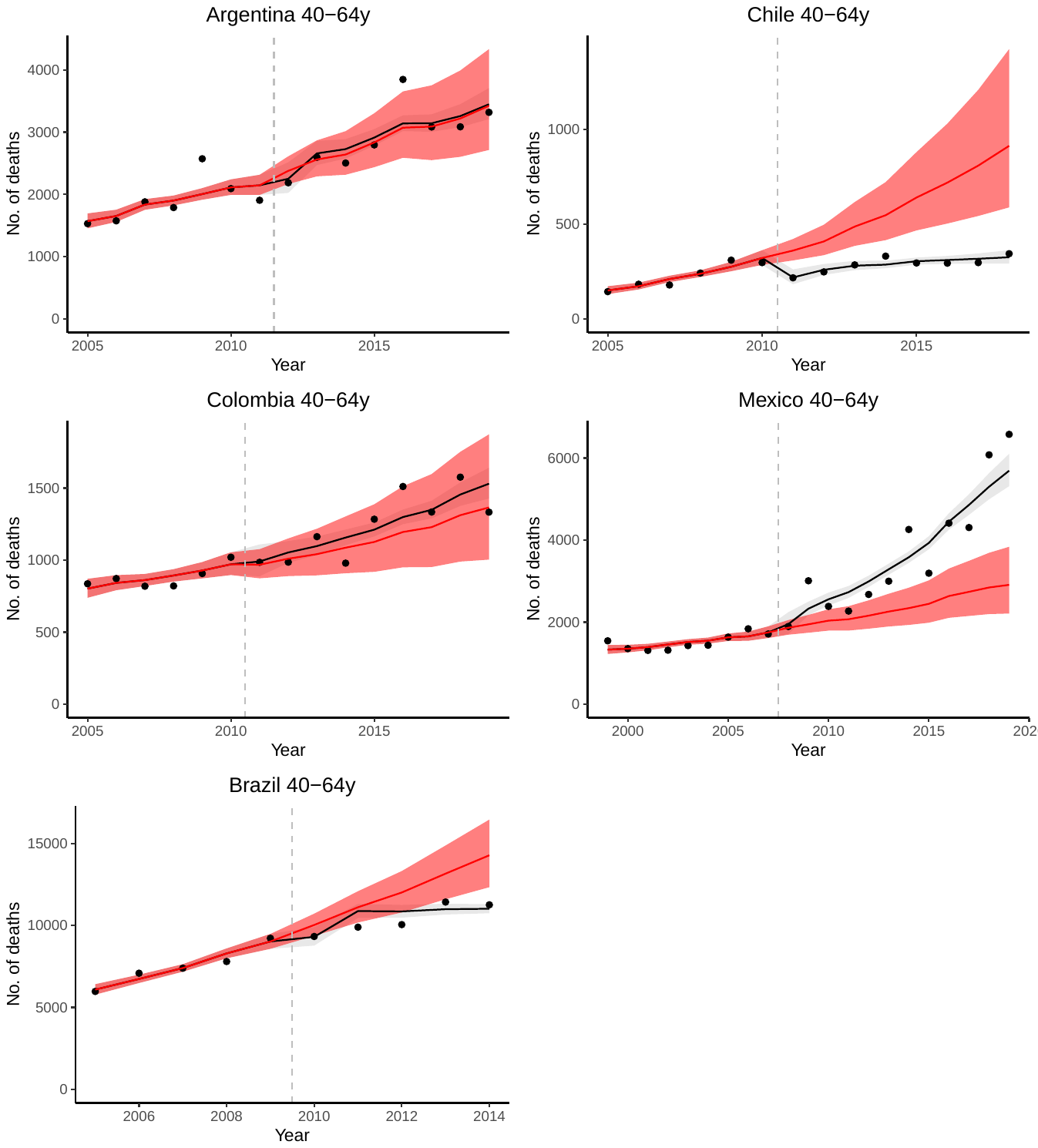

Figure S9: Annual time series for the observed and counterfactual number of pneumonia deaths among children and young adults aged 65-79 years old in 5 Latin American countries. Dots represent the observed number of pneumonia deaths (*International Classification of Diseases, Tenth Revision* codes J12–J18). Lines and gray-shaded areas represent point estimates and 95% credible intervals of the fitted pneumonia deaths from the ITS model, respectively. Red lines and shaded areas represent point estimates and 95% credible intervals of the predicted pneumonia deaths, respectively. Vertical dashed lines show the timing of pneumococcal conjugate vaccine introduction in each country.

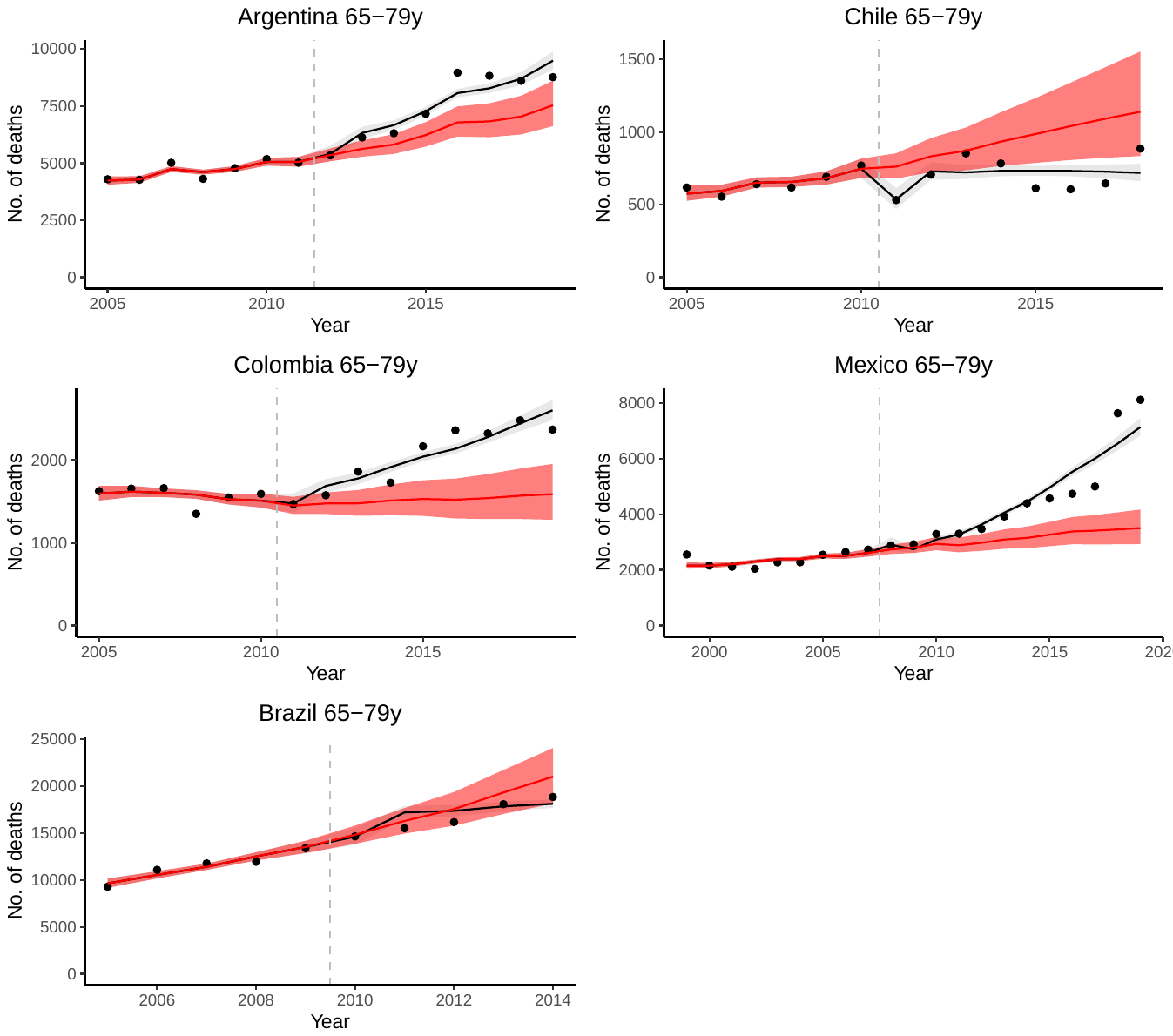

Figure S10: Annual time series for the observed and counterfactual number of pneumonia deaths among children and young adults aged 80+ years old in 5 Latin American countries. Dots represent the observed number of pneumonia deaths (*International Classification of Diseases, Tenth Revision* codes J12–J18). Lines and gray-shaded areas represent point estimates and 95% credible intervals of the fitted pneumonia deaths from the ITS model, respectively. Red lines and shaded areas represent point estimates and 95% credible intervals of the predicted pneumonia deaths, respectively. Vertical dashed lines show the timing of pneumococcal conjugate vaccine introduction in each country.

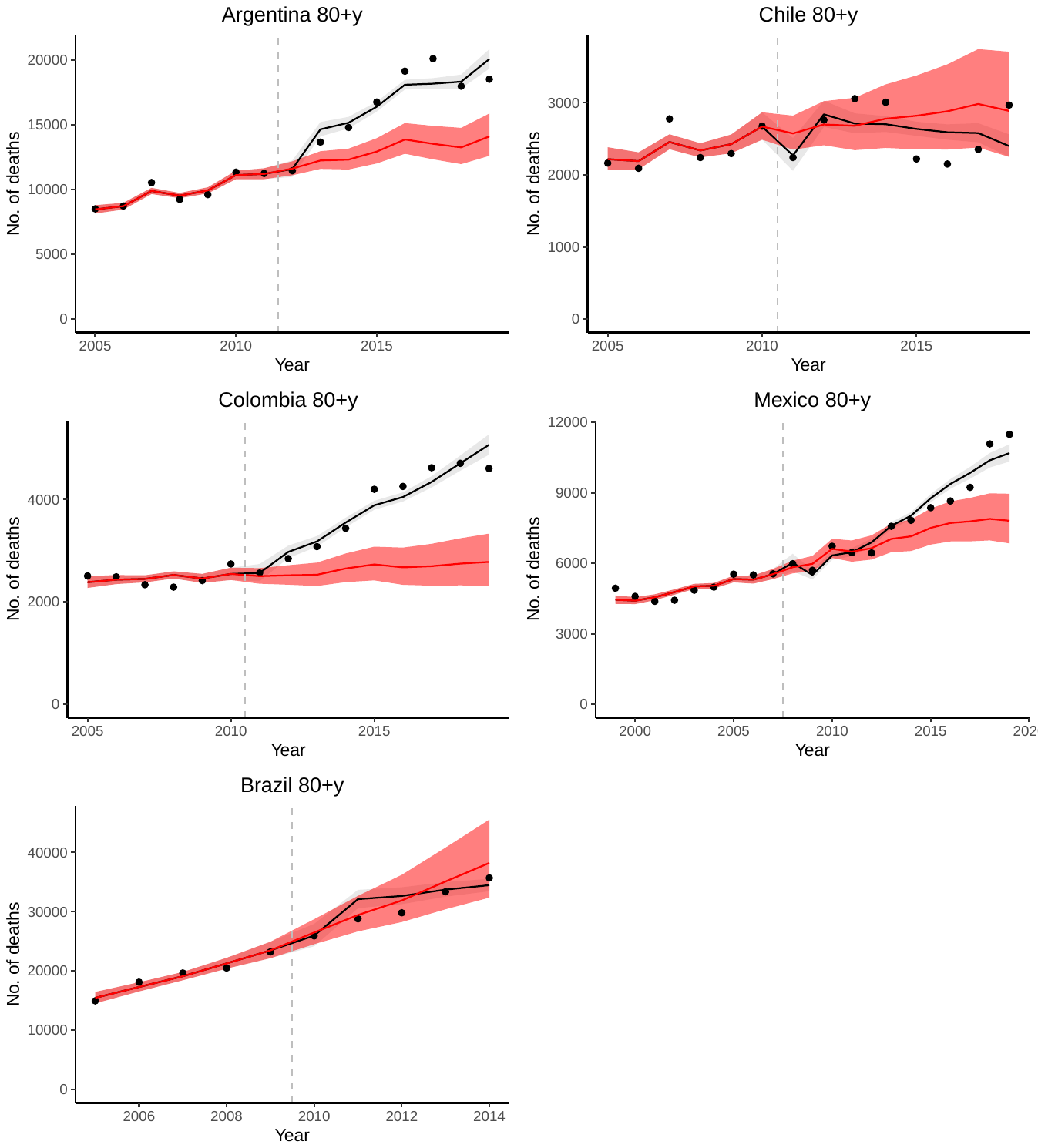

Figure S11: Estimated Impact of Pneumococcal Conjugate Vaccine in age groups 5-19 (group 3), 20-39 (group 4), 40-64 (group 5), 65-79 (group 6), 80+ (group 7) years old in Argentina (panel a), Brazil (panel c), Chile (panel e), Colombia (panel b), and Mexico (panel d) testing alternative approaches. Comparison of vaccine impact estimates from the SC, the ITS, and the SC+ITS models.

Rate ratios (RRs) were calculated by dividing the total number of observed all-cause pneumonia deaths by the total number of counterfactual all-cause pneumonia deaths duing the evaluation period.

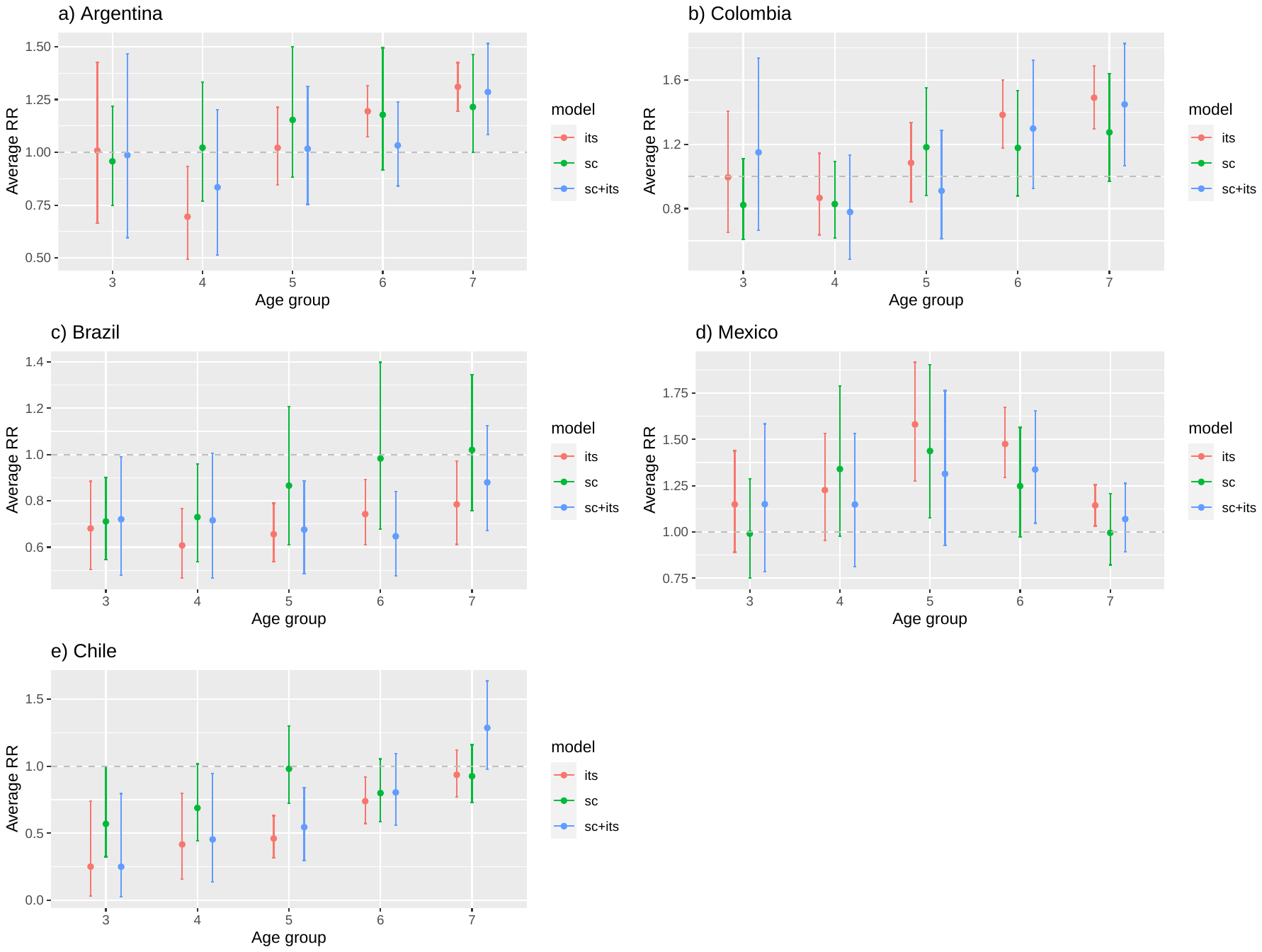

Abbreviations: SC, synthetic control; ITS, interrupted time series; RR, rate ratio.
